# Integrated Multi-omics Analysis of 105 Pediatric Germ Cell Tumors Identifies a Sphingolipid – HTRA1 – LAG3 Axis Associated with Immune Evasion in Refractory Disease

**DOI:** 10.64898/2026.05.15.26351806

**Authors:** Yixiang Song, Qingyou Zhao, Linke Yang, Hong-tao Li, Guanhua Liu, Zhongkun Guo, Siying Liu, Zhan Lei, Shuping Yang, Jingfu Wang, Manfei Liang

## Abstract

**Background:** Platinum-refractory paediatric germ cell tumours (GCTs) carry a poor prognosis, with five-year survival below 30% and no validated molecular stratification tool. The biological mechanisms underlying platinum resistance in this population remain poorly defined, limiting the development of targeted therapeutic strategies and early-warning biomarkers.

**Methods:** We performed integrated plasma multi-omics profiling in 105 pediatric GCT patients (54 refractory and 51 treatment-naïve) using data-independent acquisition proteomics, untargeted metabolomics, and exploratory lipidomics. Candidate biomarkers were validated using ELISA and spatial multiplex immunofluorescence. Predictive models were constructed using logistic regression and evaluated by ROC analysis, calibration, and decision-curve analysis.

**Results:** Multi-omics integration has revealed the coordinated dysregulation of sphingolipid metabolism, extracellular matrix remodeling, and immune checkpoint signaling in refractory diseases. Lipidomic analysis demonstrated a significant depletion of sphingolipid-associated species, including lysophosphatidylserine, lysophosphatidylethanolamine, and phosphatidylserine. Proteomic profiling identified the upregulation of LAG3 and HTRA1, which was validated by ELISA. Multiplex immunofluorescence demonstrated the spatial enrichment of exhausted CD8 + LAG3 ⁺ T cells adjacent to CK-PAN ⁺ tumor cells in refractory tumors. A plasma biomarker panel integrating LAG3, HTRA1, and AFP showed improved discrimination of refractory disease (AUC = 0.821) compared with AFP alone.

**Conclusions:** Our study identified a sphingolipid–HTRA1–LAG3 immune-evasion axis as a defining molecular feature of refractory pediatric germ cell tumors and proposed a clinically applicable plasma biomarker panel for early risk stratification.

## Introduction

Pediatric germ cell tumors (GCTs) represent a heterogeneous group of neoplasms arising from primordial germ cells, accounting for approximately 3% of all childhood malignancies ^1^. Although most patients achieve favorable outcomes with cisplatin-based chemotherapy, a significant subset (20–30%) exhibits a refractory or relapsed phenotype, which remains the primary cause of GCT-related mortality^2^. Current clinical management relies heavily on serum biomarkers such as alpha-fetoprotein (AFP) and human chorionic gonadotropin (β-hCG)^3^. However, these traditional markers are frequently inadequate for early prediction of chemoresistance, particularly in AFP-negative cases or complex histological subtypes such as mature teratomas, where tumor progression can occur despite normal marker levels^4–6^.

Recent evidence suggests that the transition to a refractory state is not merely driven by genetic mutations, but is deeply rooted in the dynamic remodeling of the tumor microenvironment (TME) and metabolic reprogramming^7 – 9^. Among proteomic drivers, high-temperature requirement A serine peptidase 1 (HTRA1) has emerged as a critical regulator of extracellular matrix integrity and growth factor signaling, and its dysregulation is linked to poor prognosis in multiple solid tumors^10 – 13^. Simultaneously, the immune landscape, characterized by exhaustion markers such as lymphocyte activation gene 3 (LAG3), plays a pivotal role in mediating immune evasion and therapy resistance in the TME^14–16^.

Beyond the proteome, metabolic shifts, particularly lipid metabolism, have been recognized as hallmarks of cancer resilience^17,18^. Sphingolipids and phospholipids, including lysophosphatidylserine (LPS) and phosphatidylserine (PS), function as more than structural components; they act as bioactive signaling molecules that modulate the IKKβ-NF-κB inflammatory axis and apoptotic thresholds^19^. Despite these insights, the synergistic interplay between the systemic proteome and the lipidome in pediatric GCTs remains largely unexplored. A holistic understanding of these cross-omic interactions is essential to identify high-risk patients who may require intensified or novel therapeutic interventions.

Despite these advances, a comprehensive multi-omics investigation integrating metabolic, proteomic, and spatial immune profiling in pediatric germ cell tumors has not been previously reported. In the present study, we performed integrated plasma multi-omics profiling using data-independent acquisition (DIA) proteomics, untargeted metabolomics, exploratory lipidomics, and spatial multiplex immunofluorescence in a cohort of 105 pediatric patients with GCT. Our analysis identified a coordinated molecular program characterized by sphingolipid metabolic dysregulation, HTRA1-mediated extracellular matrix remodeling, and LAG3-associated immune exhaustion in refractory diseases. Furthermore, we developed a plasma biomarker panel integrating LAG3, HTRA1, and AFP, which demonstrated strong predictive performance for identifying refractory tumors. These findings provide new insights into the biological mechanisms underlying treatment resistance in pediatric germ cell tumors and highlight potential biomarkers and therapeutic targets for future clinical investigation.

## Methods

### Study Design and Patient Cohort

This was a single-center, retrospective, plasma multi-omics study conducted at Shandong Cancer Hospital. Pediatric patients (age < 18 years at diagnosis or ≤ 21 years per adolescent-and-young-adult criteria) with histologically confirmed germ cell tumors (GCT) who underwent plasma collection between August 2023 and November 2025 were enrolled. Patients were classified into two groups: (1) treatment-naïve (NT), defined as patients sampled before the initiation of any systemic therapy, and (2) refractory (RF), defined as patients with disease progression, relapse, or lack of complete response after ≥ 2 prior lines of platinum-based chemotherapy. A total of 105 patients with evaluable plasma samples were included in this study (NT, n = 51; RF, n = 54). The study was approved by the Institutional Review Board of Shandong Cancer Hospital (approval number: 20250814), and written informed consent was obtained from patients and/or guardians in accordance with the Declaration of Helsinki.

### Inclusion and Exclusion Criteria

#### Inclusion criteria

(1) Histologically confirmed GCT of any anatomical site; (2) age ≤ 21 years at diagnosis; (3) adequate plasma volume for proteomics and metabolomics (≥ 300 μl); and (4) complete clinical data, including primary site, AFP, treatment history, and follow-up outcome.

#### Exclusion criteria

(1) concurrent active malignancy other than GCT; (2) plasma sample collected during active infection or severe comorbidity likely to confound the plasma proteome; (3) inadequate plasma volume; and (4) withdrawal of consent.

### Multi-Omics Profiling: Proteomics, Metabolomics and Lipidomics

Fasting venous blood (5 mL) was collected in EDTA tubes and processed within 2h via sequential centrifugation (1600 × g for 10 min followed by 16,000 × g for 10 min at 4 °C) to isolate platelet-poor plasma, which was then aliquoted and stored at −80 °C until use. Plasma proteomics and untargeted metabolomics were performed at BGI (Beijing Genomics Institute, Shenzhen, China).

Plasma proteomic profiling was performed using data-independent acquisition mass spectrometry (MS). Plasma proteins were extracted, digested with trypsin, and analyzed using a high-resolution LC – MS/MS platform. Raw spectral data were processed using DIA-based quantification software to obtain normalized protein intensity values for 105 samples, 38681 peptides and 5483 proteins were identified for downstream analysis. Differential protein expression between naïve and refractory groups was assessed using non-parametric statistical tests. Proteins with P<0.05 were considered differentially expressed. Functional enrichment analysis was performed using the Gene Ontology (GO) and KEGG pathway databases to identify the biological processes associated with treatment resistance.

Untargeted metabolomic profiling was performed using LC–mass spectrometry. Raw data were processed for peak extraction, peak alignment, and metabolite identification to generate a metabolite intensity matrix containing the peak areas and metabolite annotations. Differential metabolite analysis between the naïve and refractory groups was conducted using Mann–Whitney U tests and multivariate statistical models. Metabolic pathway enrichment analysis was performed using KEGG and MetaboAnalyst to identify the significantly altered metabolic pathways.

Targeted lipidomic profiling was performed by LC-Bio Technologies (Hangzhou, China) using LC–MS/MS. A total of 1,059 lipid species across 26 subclasses were annotated, including 430 triacylglycerols (TG), 76 phosphatidylcholines (PC), 61 sphingolipid species (SM, Cer, HexCer, and Hex2Cer), and additional lysophospholipid classes.

Differential lipid analysis between refractory and naïve tumors was performed using partial least squares discriminant analysis (PLS-DA) to calculate variable importance in projection (VIP) scores combined with the Mann–Whitney U test for statistical significance. Lipid species with VIP>1 and P<0.05 were considered significantly altered.

To evaluate global sphingolipid remodeling, the distribution of log₂ fold changes across all 61 sphingolipid species was as sessed using the Wilcoxon signed-rank test to determine whether the median log₂ fold change differed from zero. Subclass-level analyses were performed using Wilcoxon tests within the individual sphingolipid subclasses. Statistical significance was defined as P < 0.05.

### ELISA Validation

To validate the systemic concentrations of candidate biomarkers, plasma levels of HTRA1 and LAG3 were measured in the 105-patient cohort using commercially available enzyme-linked immunosorbent assay (ELISA) kits, according to the manufacturer’s protocols. All samples and standards were analyzed in duplicate. Absorbance was measured at 450 nm using a microplate reader, and concentrations were calculated using a four-parameter logistic (4PL) curve fit.

### Multiplex Immunofluorescence (mIF) and Spatial Analysis

Formalin-fixed paraffin-embedded (FFPE) tumor tissue sections (4 μ m thickness) were obtained from archival surgical specimens of six representative patients, including three AFP-positive yolk sac tumor (YST) cases from the refractory group and three AFP-negative teratoma-predominant cases from the treatment-naïve group. Multiplex immunofluorescence (mIF) staining was performed using a tyramide signal amplification–based multiplex staining system, according to the manufacturer’s protocol.

A six-marker antibody panel was designed to characterize tumor epithelial and immune cell populations, including LAG3 (OPAL 520), HTRA1 (OPAL 570), CD8 (OPAL 620), CD68 (OPAL 690), and pan-cytokeratin (CK-PAN; OPAL 480), using DAPI as a nuclear counterstain. Sequential rounds of antibody staining and fluorophore deposition were performed with microwave-based antigen retrieval between staining cycles to remove the primary antibodies while preserving the fluorescent signal.

The stained slides were scanned using an eight-channel fluorescence digital slide scanner (AF-KL-20-8, AiFang Biological, China) to generate high-resolution whole-slide images. Quantitative image analysis was performed using QuPath software (version 0.6.x). Cell detection was conducted using DAPI nuclear staining, followed by cell segmentation and classification based on marker expression. Tumor epithelial cells were defined as CK-PAN⁺ cells, whereas immune cell subsets included CD8 + cytotoxic T cells and CD68 + macrophages. Exhausted T-cells were defined as CD8 + LAG3⁺ positive cells.

For spatial analysis, cell density and nearest-neighbor distance analyses were performed to quantify the spatial relationships between immune cells and tumor epithelial cells using QuPath. In particular, the distances between CD8 + LAG3⁺ T cells and CK-PAN⁺ tumor cells were calculated to evaluate immune–tumor interactions.

### Bioinformatics and Systems Integration

Pathway activity scores were estimated using single-sample gene set enrichment analysis (ssGSEA). Kinase – substrate regulatory networks were constructed to identify signaling pathways associated with differential protein expression. Integrated multi-omics analysis was performed by combining protein–lipid correlation modules with kinase signaling networks to identify key regulatory pathways.

### Statistical Analysis and Predictive Modeling

Continuous variables were compared using Student’s t-test or the Mann-Whitney U test. Event-free survival (EFS) was defined as the time from sample collection to disease progression, relapse, or death and was estimated using the Kaplan-Meier method. Multivariate Cox regression was performed to identify independent prognostic factors, adjusting for AFP level and clinical stage. Based on the independent predictors, a prognostic nomogram was developed and internally validated using Harrell’s C-index and 1,000 bootstrap resamples. Decision Curve Analysis (DCA) was conducted to evaluate the net clinical benefit and practical utility of the model. Correlation analysis was performed using Spearman’s rank correlation test. All statistical analyses were conducted using R software (version 4.3.0) and GraphPad Prism (version 10.0). Two-sided P values < 0.05 were considered statistically significant.

## Results

### Clinical characteristics of the study cohort

A total of 105 pediatric patients with germ cell tumor (pGCT) were included in this study, comprising 51 treatment-naïve and 54 treatment-refractory cases. The baseline clinical characteristics of the patients are summarized in Table 1. There were no significant differences in age (median 13.0 vs. 16.0 years, P = 0.251) or sex distribution (P = 0.521) between the two groups. Similarly, initial serum AFP levels at the time of diagnosis were comparable (P = 0.178).

**Table 1.**
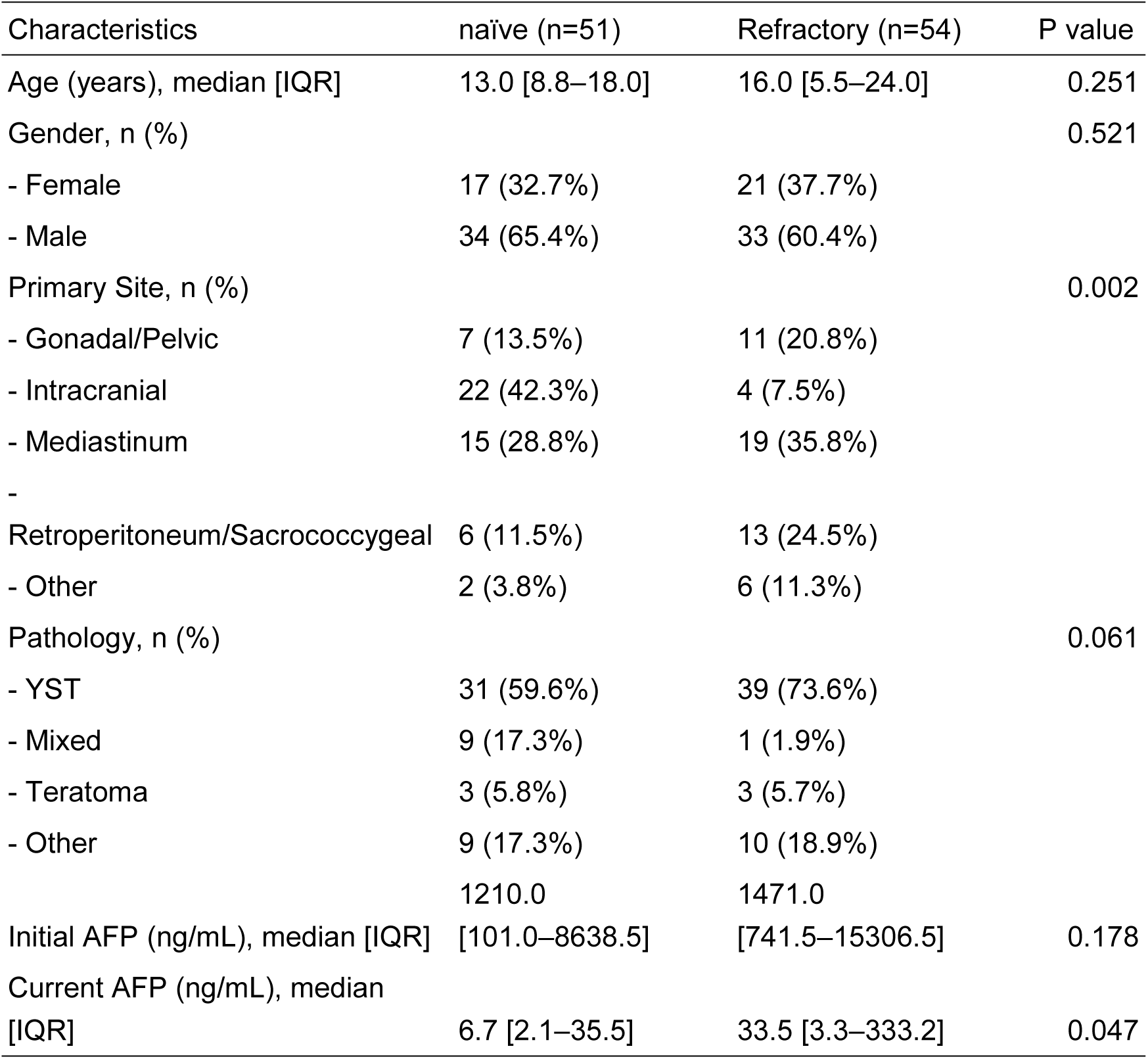
Baseline clinical characteristics of patients with GCTs.

However, the distribution of primary tumor sites differed significantly between the groups (P=0.002). Intracranial tumors were more frequent in the treatment-naïve cohort (42.3% vs. 7.5%), whereas mediastinal and retroperitoneal/sacrococcygeal tumors were more common in refractory patients.

Although the pathological subtypes did not reach statistical significance (P=0.061), yolk sac tumor (YST) was more prevalent in refractory cases (73.6% vs. 59.6%). Notably, AFP levels at the time of sampling were significantly elevated in refractory patients compared with treatment-naïve patients (median 33.5 vs. 6.7 ng/mL, P=0.047), consistent with persistent disease activity.

### Global proteomic and metabolomic profiling reveals coordinated immune and metabolic alterations in refractory pediatric GCTs

To comprehensively characterize the molecular alterations underlying therapy resistance in pediatric germ cell tumors (GCTs), we designed a multi-omics workflow to profile plasma samples from 105 patients with plasma-derived GCTs (pGCTs), including 51 treatment-naïve and 54 refractory cases (Figure 1A). First, plasma samples were subjected to DIA-based proteomics, complemented by metabolomics and lipidomics profiling. Quality control (QC) assessments confirmed high data reproducibility and robust normalization across the cohorts (Supplementary Fig. S1A). In total, 5483 plasma proteins were quantified (Supplementary Table S1). Principal component analysis (PCA) revealed distinct segregation between refractory and treatment-naïve patients, indicating that therapeutic resistance was associated with specific systemic proteomic signatures (Figure 1B).

**Figure 1.**
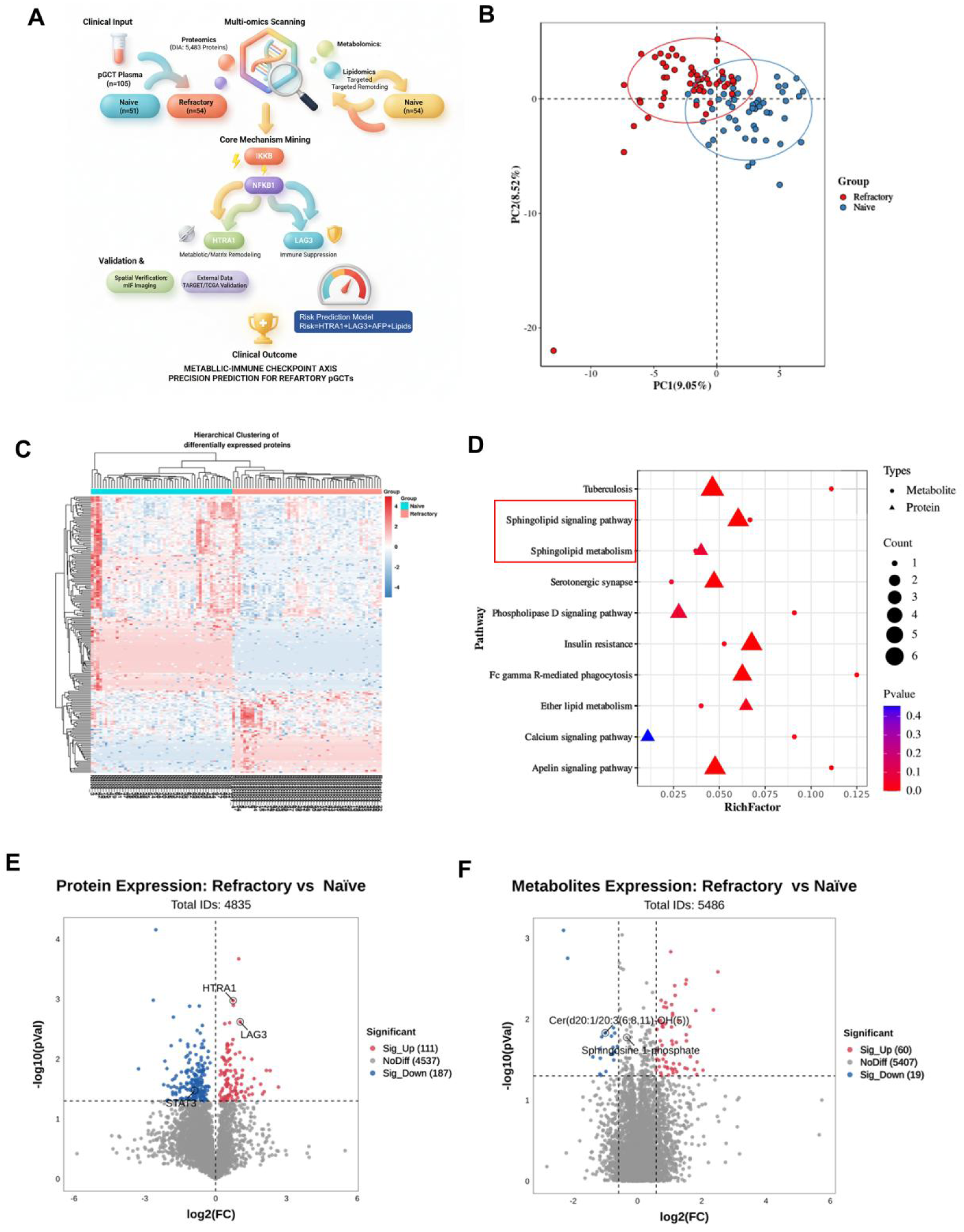
Integrated plasma proteomic and metabolomic profiling identifies a distinct molecular landscape in refractory pediatric germ cell tumors. (A) Study workflow showing plasma-based DIA proteomics and untargeted metabolomic profiling in 105 pediatric patients with GCT (B) Principal component analysis (PCA) of plasma proteomics demonstrating partial separation between treatment-naïve and treatment-refractory patients. (C) Hierarchical clustering heatmap of the differentially expressed proteins. (D) Functional enrichment analysis highlighting the immune-related and sphingolipid metabolic pathways. (E) Volcano plot of differentially expressed plasma proteins between refractory and treatment-naïve groups. Significantly dysregulated proteins (FDR-adjusted P < 0.05) are highlighted. (F) Volcano plot of untargeted metabolomic analysis showing significantly altered metabolites in refractory and treatment-naïve patients.

Differential expression analysis identified 202 proteins that were significantly dysregulated between the two groups (FDR<0.05, fold-change >1.5; Supplementary Table S2). Hierarchical clustering of these differentially expressed proteins (DEPs) effectively categorized patients based on their clinical responses, reinforcing the biological robustness of the refractory-associated proteomic signature (Figure. 1C).

To elucidate the functional implications of these alterations, we performed integrative pathway enrichment analysis. This multi-layered approach identified sphingolipid metabolism and sphingolipid signaling pathway as the most significantly enriched processes in refractory patients (P<0.001), with synergistic contributions from both proteomic and metabolomic datasets (Figure. 1D; full enrichment results are provided in Supplementary Table S3). Notably, several key regulators of immune evasion and microenvironmental remodeling were among the most significantly upregulated proteins in the refractory cohort, including the immune checkpoint molecule LAG3 and the extracellular matrix (ECM) serine protease HTRA1 (Figure 1E, FDR-adjusted P<0.05; Supplementary Table S4). Concurrently, untargeted metabolomics corroborated this widespread metabolic reprogramming, specifically validating the dysregulation of sphingolipid metabolism through the significant alteration of various ceramide and sphingosine species (Figure 1F, FDR-adjusted P < 0.05; Supplementary Table S5). Collectively, these data suggest a coordinated metabolic-immune axis that characterizes the refractory phenotype in pediatric GCTs.

### Integrated plasma lipidomic and proteomic profiling identifies biomarkers associated with refractory pediatric germ cell tumors

Pathway analysis (Figure 1) identified sphingolipid signaling as a central dysregulated process in the refractory group. To further investigate the molecular components underlying dysregulation of this pathway, we performed plasma lipidomic profiling.

Unsupervised clustering of the top differentially expressed proteins demonstrated clear separation between the refractory and naïve groups (Figure. 2A), indicating that distinct systemic protein signatures were associated with treatment resistance. Consistent with the pathway enrichment results, several phospholipid- and sphingolipid-related lipid species showed significant alterations in refractory tumors. Lysophosphatidylserine (LPS), lysophosphatidylethanolamine (LPE), and phosphatidylserine (PS) levels were significantly higher in refractory than in naïve patients (Figure. 2B-D), indicating substantial remodeling of the membrane lipid metabolism.

**Figure 2.**
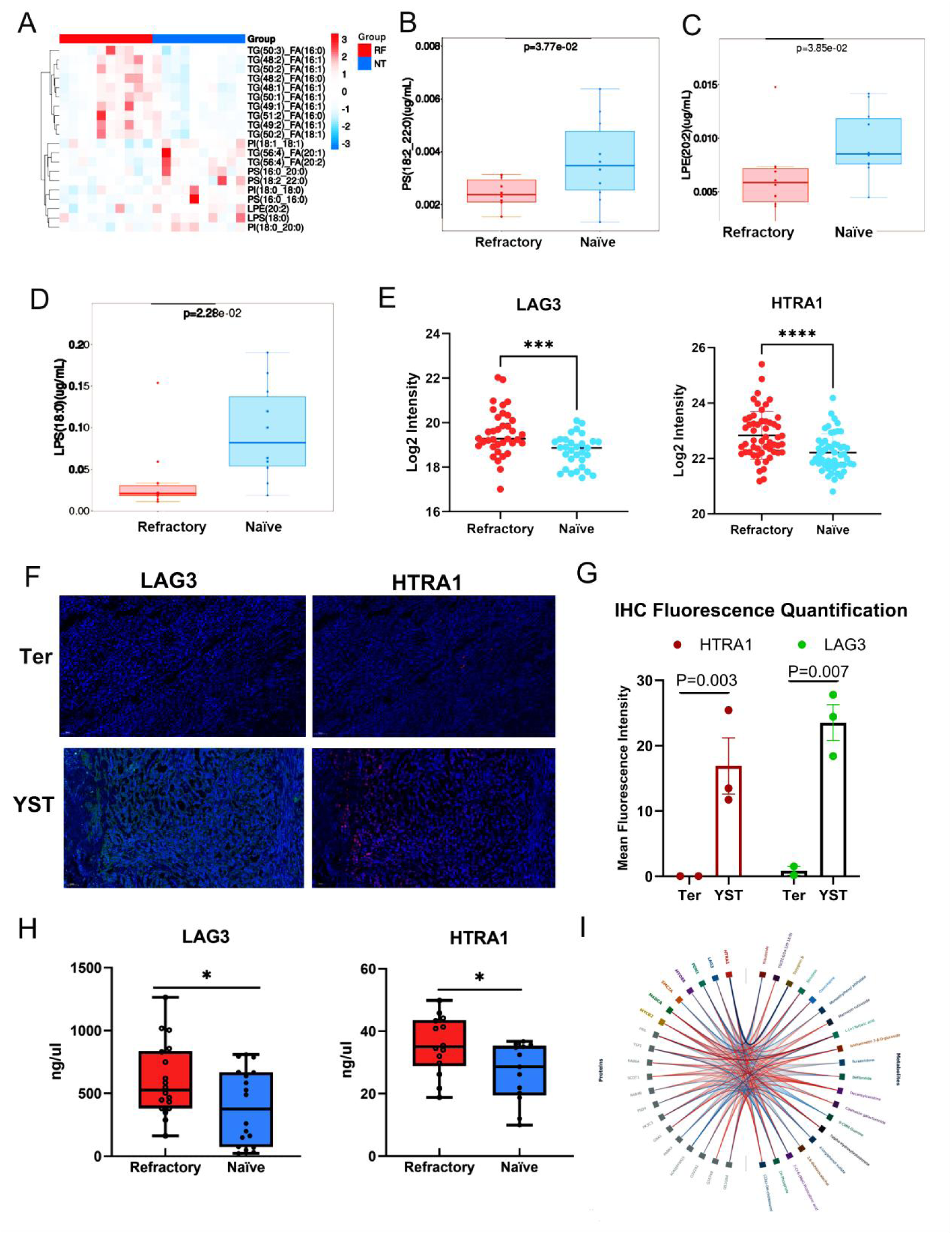
Integrated plasma lipidomic and proteomic profiling identifies biomarkers associated with refractory pediatric germ cell tumors. (A) Heatmap showing representative differentially expressed lipid species between refractory (RF) and naïve (NT) pediatric germ cell tumors. Red indicates relatively increased abundance, whereas blue indicates decreased abundance. Differential lipids were mainly enriched in phospholipid-related subclasses, including phosphatidylserine (PS), lysophosphatidylethanolamine (LPE), and lysophosphatidylserine (LPS). (B – D) Boxplots showing quantitative differences in representative lipid metabolites between RF and NT groups, including PS(18:2_22:0) (B), LPE(20:2) (C), and LPS(18:0) (D). P values were calculated using the Mann–Whitney U test. (E) Differential expression analysis of plasma LAG3 and HTRA1 protein levels between RF and NT groups based on DIA proteomics. Scatter plots show log2-transformed protein intensities. Horizontal lines indicate mean values. Statistical significance was determined using the Student’s t-test. (F) Representative multiplex immunofluorescence (mIF) images of LAG3 and HTRA1 expression in teratoma-predominant (Ter) and yolk sac tumor (YST) specimens. (G) Quantification of mIF fluorescence intensity for HTRA1 and LAG3 in Ter and YST tissues. Data are presented as mean ± SD. (H) ELISA validation of plasma LAG3 and HTRA1 levels in RF and NT patients. Both proteins were significantly elevated in refractory tumors. (I) Integrated protein – metabolite interaction network showing correlations between differentially expressed proteins and lipid metabolites. Red and blue connecting lines indicate positive and negative correlations, respectively.

This redistribution of lipid classes is consistent with the sphingolipid pathway perturbation identified at the metabolomic level, suggesting a broader dysregulation of lipid homeostasis in platinum-refractory diseases. The full lipid species list and statistical parameters are provided in Supplementary Table S6, and the top20 variable contributors are shown in Supplementary Fig. S1B. Although these lipid species are canonical components of glycerophospholipid metabolism, they are metabolically connected to sphingolipid biology through shared biosynthetic intermediates and regulatory lipid flux.

Consistent with the metabolomic findings, DIA proteomics revealed significant upregulation of the immune exhaustion marker LAG3 and the extracellular matrix-associated protease HTRA1 in refractory tumors (Figure 2E). These observations were further validated at the tissue level by multiplex immunofluorescence (mIF). Representative staining demonstrated markedly stronger LAG3 and HTRA1 fluorescence signals in yolk sac tumor (YST) tissues compared with teratoma-predominant specimens (Figure 2F). Quantitative fluorescence analysis confirmed significantly increased expression of both markers in YST samples (Figure 2G).

To validate the circulating protein alterations, ELISA assays were performed in an independent plasma cohort. Both LAG3 and HTRA1 concentrations were significantly elevated in refractory patients relative to naïve cases (Figure 2H), supporting the reproducibility of the proteomic findings.

Integrated protein–metabolite network analysis further demonstrated extensive correlations between altered lipid metabolites and differentially expressed proteins (Figure 2I). Notably, phospholipid-associated metabolites showed coordinated associations with immune- and extracellular matrix-related proteins, suggesting that lipid metabolic remodeling may contribute to the establishment of an immunosuppressive tumor microenvironment in refractory pGCT.

Subgroup analyses further demonstrated that the expression levels of LAG3 and HTRA1 were not significantly associated with AFP status or tumor histology (Supplementary Fig. S1 C–F). This lack of subgroup-specific variation underscores the LAG3-HTRA1 axis is a universal hallmark of the refractory phenotype that transcends conventional clinical parameters, offering a robust and independent tool for identifying high-risk patients who might be missed by traditional histology-based screening.

To further investigate the molecular synergy driving tumor progression, we performed cross-omics integration of proteomic and metabolomic profiles. The global correlation architecture revealed a highly connected molecular network centered around the LAG3-HTRA1 axis (Figure 2 I).

Collectively, these findings identify LAG3 and HTRA1 as key circulating biomarkers associated with refractory pediatric germ cell tumors, highlighting their potential involvement in immune dysregulation and tumor microenvironment remodeling. Notably, the coordinated upregulation of the LAG3 – HTRA1 axis was consistently observed across refractory cases irrespective of conventional clinicopathological features, suggesting that this axis represents a molecular hallmark of treatment-resistant disease. These results further support its potential utility as a robust biomarker framework for improving risk stratification and identifying high-risk patients beyond traditional histology- or AFP-based assessment.

### Integrated lipid–NF-κB regulatory network linking sphingolipid metabolism to immune checkpoint activation

To decipher the systemic molecular interactions driving tumor refractoriness, we constructed an integrated multi-omics regulatory network. Kinase regulatory analysis identified STAT3 and NFKB1 as primary hubs within the proteomic landscape, suggesting their central role in mediating treatment resistance (Figure 3A). Interestingly, while core components of the IKK/NF-κB complex were down-regulated at the protein level (Figure 3B), pathway enrichment analysis revealed a significant activation of the broader Sphingolipid signaling pathway in the refractory cohort. Functional mapping of this pathway indicated that metabolic dysregulation, specifically involving S1P, LPS, and LPE, was intrinsically coupled with NF- κ B-dependent pro-survival signaling (Figure 3C). This metabolic-signaling coupling was further supported by cross-omics correlation matrices, which showed that the stromal-remodeling protease HTRA1 and the immune-checkpoint LAG3 are linked to specific lipid signatures (Figure 3D).

**Figure 3.**
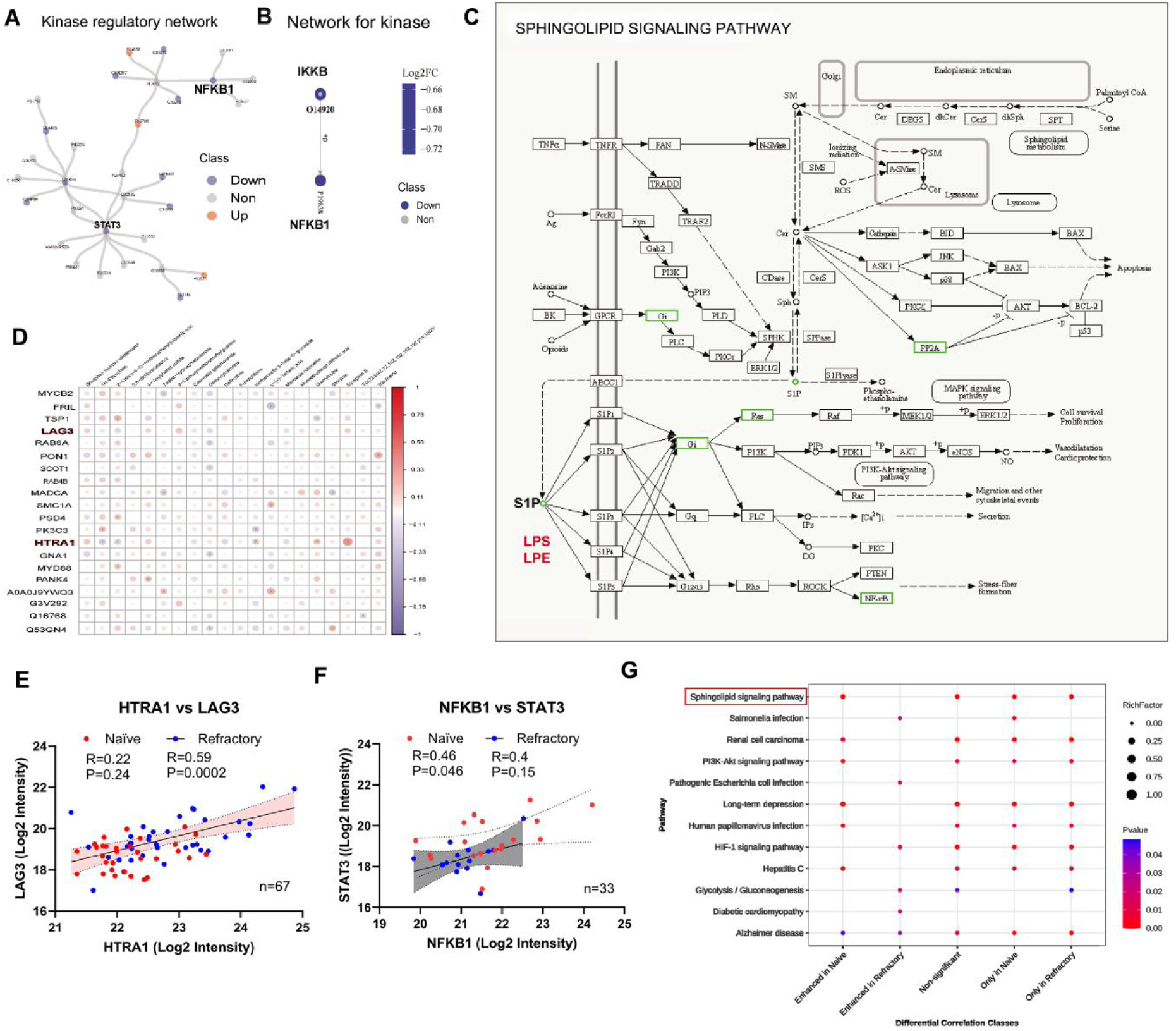
Integrated network analysis reveals dysregulated sphingolipid–NF-κB signaling in refractory pediatric GCTs. (A) Kinase regulatory network constructed from differentially expressed genes (DEGs) between refractory and naïve pediatric GCTs. Red nodes represent upregulated kinases, blue nodes represent downregulated kinases, and gray nodes represent non-significantly changed molecules. NFKB1 and STAT3 were identified as core hub kinases with the highest connectivity in the network.(B) Hub kinase interaction network highlighting the key regulatory axis of IKBKB-O14920-PI9638-NFKB1, with node color indicating log₂(fold change) (refractory vs. naïve) and node size representing connectivity degree.(C) Correlation heatmap of hub kinases with hallmark signaling pathways. The size and color of dots represent the strength and significance of Spearman correlation, respectively. NFKB1 showed the strongest negative correlation with multiple immune-related and oncogenic pathways, including NF-κB signaling, HIF-1 signaling, and PD-L1/PD-1 checkpoint pathway.(D) Schematic diagram of the NF-κB signaling pathway, with green boxes marking downregulated molecules (including NFKB1) in refractory GCTs, confirming the suppressed activation of NF-κB pathway in drug-resistant tumors. (E) Correlation between HTRA1 and LAG3 expression in the combined cohort (n=67). Pearson correlation analysis was performed separately in the naïve (red dots, R=0.22, P=0.24) and refractory (blue dots, R=0.59, P=0.0002) groups. (B) (F) Correlation between NFKB1 and STAT3 expression in the combined cohort (n=33). Pearson correlation analysis was performed separately in the naïve (red dots, R=0.46, P=0.046) and refractory (blue dots, R=0.40, P=0.15) groups. (C) (G) Differential correlation analysis of signaling pathways between naïve and refractory patients. The bubble plot displays pathways with significantly different correlation patterns, including the sphingolipid signaling pathway (highlighted in red), with color intensity indicating P-values and bubble size representing the RichFactor.

To characterise the protein-level co-expression relationships underlying the refractory plasma proteome, we examined pairwise Pearson correlations between key biomarker proteins. HTRA1 and LAG3 demonstrated a significant positive co-expression relationship (R = 0.59, P = 0.0002, n = 67; Figure 3E).The positive co-expression of HTRA1 and LAG3 is mechanistically coherent: both proteins are downstream effectors of the NF- κ B suppression cascade, with NF- κ B inactivation simultaneously de-repressing HTRA1 transcription and permitting LAG3 upregulation on tumour-infiltrating T cells ^14,20^. Their correlated elevation in refractory plasma thus reflects a unified molecular resistance programme rather than two independent events.

In treatment-naïve patients, NFKB1 and STAT3 showed a statistically significant positive correlation (R = 0.46, P = 0.046, n = 33), indicating that the NF- κ B – STAT3 transcriptional co-regulatory axis is functionally intact in the treatment-naïve plasma proteome. In contrast, in platinum-refractory patients, this NFKB1 – STAT3 coupling was substantially attenuated and statistically non-significant (R = 0.40, P = 0.15, n = 33; Figure 3F). Although the correlation coefficients in the two groups are numerically similar, the loss of statistical significance in the larger refractory group — despite greater statistical power — indicates a genuine reduction in co-expression fidelity rather than a power artefact.

Functionally, this NF- κ B – STAT3 uncoupling in refractory disease has two convergent consequences: de-repression of HTRA1 transcription (as NF- κ B normally represses the HTRA1 promoter), and impairment of MHC class II and co-stimulatory molecule expression driven by STAT3, thereby creating a tumour microenvironment permissive to LAG3-mediated T-cell exhaustion. Together with the HTRA1 – LAG3 co-expression (Figure 3E), the NFKB1 – STAT3 uncoupling in refractory GCT provides protein-level co-expression evidence for the sphingolipid– NF- κ B– HTRA1/LAG3 regulatory cascade that constitutes the mechanistic backbone of platinum resistance in this cohort.

To contextualise these protein co-expression findings within broader pathway biology, we performed differential correlation analysis using the DGCA framework1 on WGCNA-derived sub-network, the co-expression module with the highest sphingolipid gene representation (Figure 3G). KEGG pathway enrichment of the differentially correlated protein pairs identified Sphingolipid signalling pathway as the most prominently enriched pathway, present across multiple differential correlation classes — including Enhanced in Naïve and Only in Refractory — indicating that sphingolipid co-expression relationships are fundamentally rewired between treatment groups. This finding directly corroborates the lipidomics evidence of global sphingolipid depletion in refractory plasma (Wilcoxon P = 0.0012; Figure 2D) and the joint proteomics–metabolomics pathway enrichment analysis identifying Sphingolipid signalling (p.adj = 0.0038) and Sphingolipid metabolism (p.adj = 0.0071) as co-enriched pathways (Figure 1D), providing orthogonal transcriptome-surrogate evidence of sphingolipid pathway dysregulation at the network co-expression level.

Additional significantly enriched pathways included PI3K–Akt signalling, HIF-1 signalling, and Glycolysis/Gluconeogenesis, consistent with a coordinated metabolic adaptation programme in refractory paediatric GCT encompassing aerobic glycolysis, hypoxia response, and growth factor signalling.

The functional consequences of these alterations were further assessed using single-sample gene set enrichment analysis (ssGSEA). Refractory tumors exhibit increased activity in immune signaling, extracellular matrix (ECM) remodeling, lipid metabolism, and inflammatory pathways (Supplementary Fig. S2A). Correlation analyses showed that LAG3 expression was strongly associated with immune pathway activity, whereas HTRA1 was linked to ECM remodeling, suggesting that these factors act as key intermediates connecting metabolic dysregulation with tumor microenvironment remodeling (Supplementary Fig. S2B–D).

Multi-omics integration further identified a coordinated regulatory network centered on HTRA1 and LAG3 (Supplementary Fig. S2E). Lysophospholipid species, including LPS, LPE, and PS, are closely associated with immune activity and ECM remodeling scores, supporting a model in which lipid metabolic perturbations propagate through HTRA1-dependent signaling to modulate immune checkpoint activation. Protein interaction analysis further confirmed HTRA1 is a central hub linking lipid metabolism and immune signaling pathways (Supplementary Fig. S2F).

Collectively, these findings reveal that refractory pediatric GCTs are characterized by coordinated disruption of the sphingolipid metabolism–NF-κB signaling axis, leading to downstream activation of ECM remodeling and immune checkpoint pathways. This integrated lipid – immune regulatory mechanism provides a mechanistic framework for immune dysfunction and therapeutic resistance in refractory disease.

### Spatial immune microenvironment characterized by HTRA1–LAG3 signaling

Given that lipidomic and phosphoproteomic analyses (Figure 3) indicated activation of sphingolipid-associated inflammatory signaling and NF-κB pathways, we hypothesized that these molecular alterations might be accompanied by coordinated remodeling of the tumor microenvironment (TME). To investigate whether the lipid–immune signaling axis identified in the multi-omics analysis was reflected at the spatial level, we performed six-plex multiplex immunofluorescence (mIF) staining to characterize the spatial relationships among HTRA1 expression, tumor epithelial cells, and immune cell populations.

Whole-section mIF imaging revealed striking contrast between the tumor subtypes (Figure 4A). In refractory yolk sac tumor (YST01) specimens, abundant immune infiltration was observed throughout the tumor parenchyma, with strong HTRA1 signals co-occurring in regions enriched with LAG3 + and CD68 + macrophages. In contrast, mature teratoma sections (Ter01) displayed markedly weaker fluorescence signals across immune markers, with immune cells largely restricted to sparse stromal regions, consistent with the well-recognized immunologically “cold” phenotype of teratomas.

**Figure 4.**
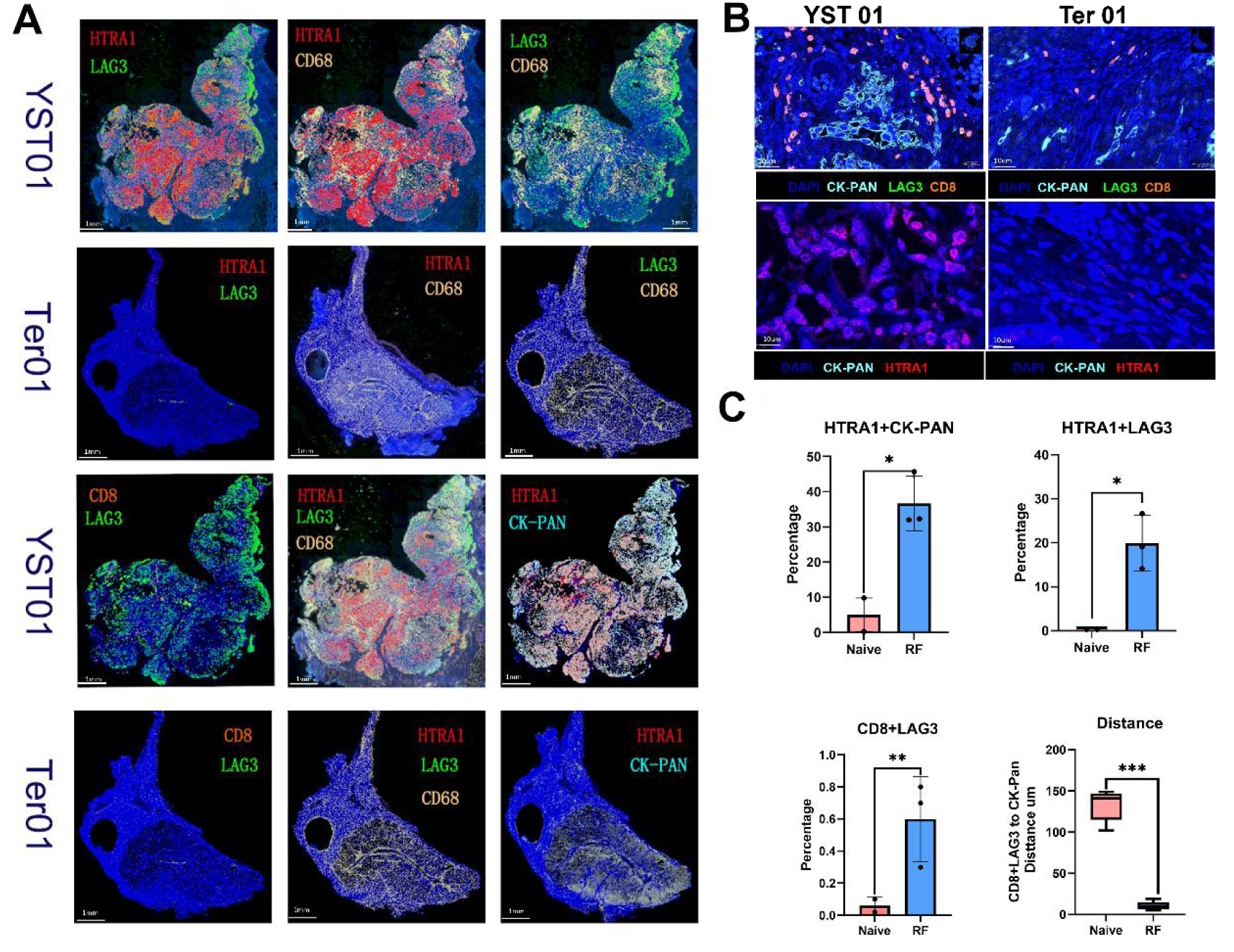
Spatial immune microenvironment of refractory pediatric germ cell tumors revealed by multiplex immunofluorescence. (A) Whole-section multiplex immunofluorescence (mIF) imaging showing the spatial distribution of HTRA1, LAG3, CD8, CD68, and CK-PAN in refractory yolk sac tumor (YST01) and mature teratoma (Ter01) specimens. (B) High-magnification views illustrating the spatial relationship between immune cells and tumor epithelial compartments. In YST tissues, LAG3⁺ immune cells and CD8⁺ T cells accumulate in close proximity to CK-PAN ⁺ tumor regions, whereas teratoma samples show sparse immune infiltration. HTRA1 expression is predominantly localized within tumor epithelial compartments. (C) Quantitative spatial analysis demonstrating increased proportions of HTRA1⁺CK-PAN ⁺ cells, elevated CD8⁺LAG3⁺ exhausted T cells, and reduced spatial distance between CD8⁺LAG3⁺ T cells and tumor epithelial cells in refractory tumors compared with naïve tumors.

High-magnification imaging further demonstrated that LAG3 ⁺ immune cells were preferentially localized in close proximity to the CK-PAN⁺ tumor epithelial compartments in YST tissues (Figure 4B). In contrast, the mature teratoma specimen exhibited minimal CD8 ⁺ or LAG3 ⁺ immune cell infiltration, indicating limited immune–tumor interactions. Consistent with proteomic findings, HTRA1 immunoreactivity was strongly enriched in the CK-PAN ⁺ tumor epithelial regions. Notably, HTRA1 staining in YST specimens was predominantly localized to the nuclei of tumor cells, suggesting that tumor epithelial cells represent a major source of HTRA1 expression within the tumor microenvironment.

Quantitative spatial analysis confirmed these observations (Figure 4C). The proportions of HTRA1 ⁺ CK-PAN ⁺ tumor cells and HTRA1 ⁺ LAG3 ⁺ cellular proximity events were both significantly increased in refractory tumors compared with treatment-naïve tumors. In parallel, the frequency of CD8 + LAG3 ⁺ exhausted cytotoxic T-cells was elevated in refractory samples, suggesting the presence of a dysfunctional T cell phenotype. Importantly, spatial distance analysis revealed that CD8 + LAG3⁺ T cells were positioned significantly closer to CK-PAN⁺ tumor cells in refractory tumors than in naïve tumors (P < 0.001). This spatial configuration indicates that exhausted T cells accumulate in close proximity to tumor cells, but are likely to remain functionally suppressed, which is consistent with localized immune exhaustion within the tumor niche.

Taken together, these findings demonstrate that refractory pediatric GCTs harbor a spatially organized immunosuppressive microenvironment characterized by HTRA1 upregulation, accumulation of LAG3⁺ immune cells, and increased CD8 + T-cell exhaustion. These spatial alterations are consistent with the lipid-driven inflammatory signaling and NF-κB pathway activation identified in upstream multi-omics analyses (Figure 3), thereby linking metabolic reprogramming with immune microenvironment remodeling in refractory diseases.

### Prognostic significance and predictive modeling of multi-omics biomarkers in pediatric germ cell tumors

To evaluate the clinical relevance and predictive value of the identified biomarkers, we assessed their association with patient outcomes, and constructed a multi-marker predictive model for refractory pediatric germ cell tumors.

Univariate Cox regression analysis demonstrated that elevated circulating levels of LAG3 and HTRA1, together with increased levels of lipid biomarkers, were significantly associated with refractory disease (Figure 5A). Among these variables, LAG3 and HTRA1 exhibited the strongest effect sizes, indicating their potential roles as independent predictors of treatment resistance.

**Figure 5.**
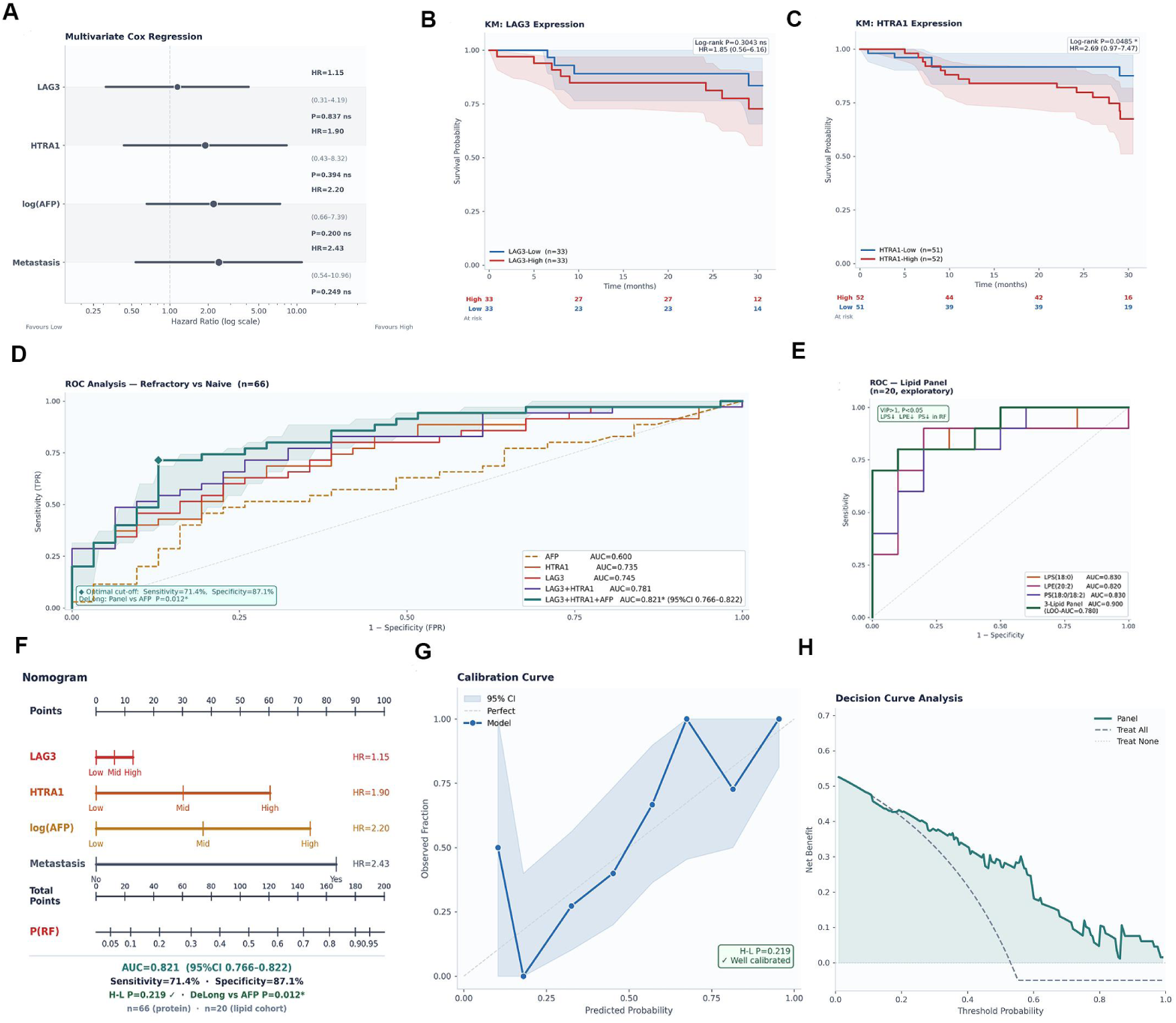
Prognostic significance and predictive modeling of multi-omics biomarkers in pediatric germ cell tumors. (A) Forest plot showing hazard ratios from Cox regression analysis of candidate biomarkers. (B–C) Kaplan–Meier survival curves stratified by circulating LAG3 and HTRA1 expression levels. Patients with higher expression exhibited significantly worse event-free survival. (D) Receiver operating characteristic (ROC) curves comparing the predictive performance of the integrated biomarker panel and individual markers. (E) ROC analysis of representative lipid metabolites demonstrating their diagnostic performance for refractory disease. (F) Nomogram incorporating LAG3, HTRA1, and lipid biomarkers for individualized prediction of refractory pediatric germ cell tumors. (G) Calibration curve evaluating the agreement between predicted and observed probabilities. (H) Decision curve analysis (DCA) assessing the clinical net benefit of the predictive model.

Kaplan–Meier survival analysis further confirmed the prognostic relevance of these markers. Patients with high LAG3 expression showed significantly reduced event-free survival compared with those with low expression (Figure 5B). Similarly, elevated HTRA1 levels were associated with poorer clinical outcomes (Figure 5C), suggesting that activation of the HTRA1–LAG3 axis may contribute to aggressive tumor behavior.

Receiver operating characteristic (ROC) analysis demonstrated that the combined biomarker panel exhibited strong predictive performance for refractory disease. The integrated multi-omics model achieved the highest discriminative accuracy compared with the individual markers (Figure 5D). Among the lipid metabolites, several species, including LPS, PS, and LPE, also showed moderate predictive ability (Figure 5E), which is consistent with the lipid metabolic alterations identified in earlier analyses.

Based on these independent predictors, we constructed a nomogram incorporating LAG3, HTRA1, and key lipid biomarkers to estimate the probability of refractory disease (Figure 5F). Each variable contributed proportionally to the overall risk score, enabling individualized prediction of treatment resistance.

The calibration curve demonstrated a good agreement between the predicted and observed probabilities of refractory disease (Figure 5G). Decision curve analysis further indicated that the model provided a positive net clinical benefit across a wide range of threshold probabilities (Figure 5H), supporting its potential utility for clinical risk stratification.

### Integrated working model of lipid–immune signaling driving platinum resistance in pediatric germ cell tumors

Multi-omics analyses revealed metabolic dysregulation characterized by altered sphingolipid metabolism and depletion of lysophospholipids (LPS and LPE), leading to perturbation of NF-κB signaling pathways. Circulating biomarkers, including LAG3 and HTRA1 have been identified as independent predictors of inferior event-free survival. Within the tumor microenvironment, refractory tumor cells exhibit nuclear accumulation of HTRA1, which may promote extracellular matrix (ECM) remodeling and reshape the immune landscape. These changes were associated with increased infiltration of LAG3 ⁺ immune cells, including exhausted CD8 + T cells and CD68 + macrophages, forming an immunosuppressive niche that facilitates tumor immune evasion. Collectively, this lipid–HTRA1–LAG3 signaling axis may contribute to the development of platinum resistance in pediatric germ cell tumors (Figure 6).

**Figure 6.**
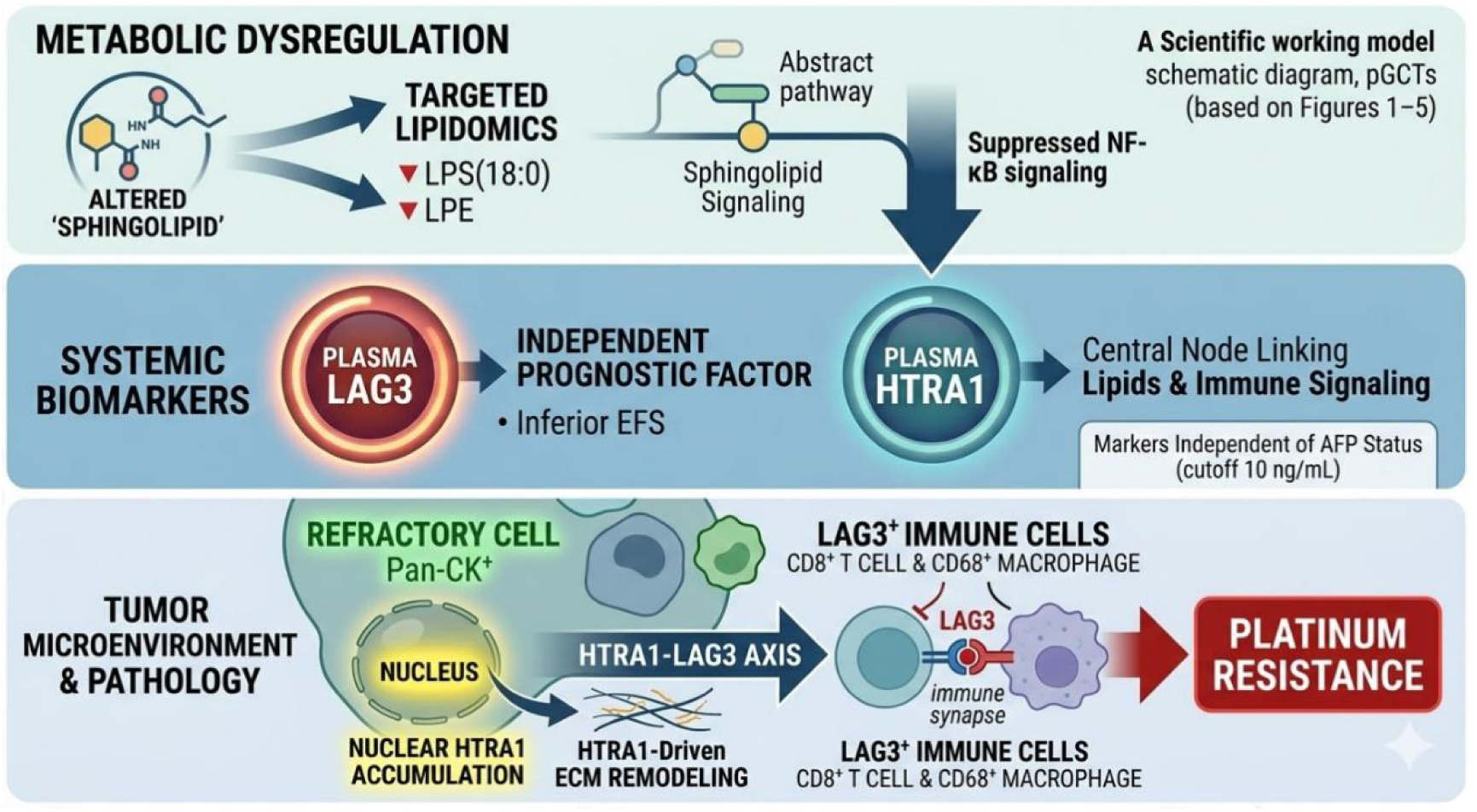
Proposed working model of lipid–immune regulatory axis driving therapeutic resistance in pediatric GCTs.

## Discussion

Platinum-refractory pediatric germ cell tumors (GCTs) remain a major clinical challenge with limited effective second-line therapies and poor survival outcomes. In this study, we performed integrated plasma multi-omics profiling of 105 pediatric patients with GCT (54 refractory and 51 treatment-naïve) using DIA proteomics, metabolomics, lipidomics, ELISA validation, and multiplex immunofluorescence. Our analyses converged on three interconnected biological programs defining refractory disease: LAG3-mediated immune exhaustion, HTRA1-driven extracellular matrix (ECM) remodelling, and global sphingolipid metabolic depletion. These pathways collectively form a coherent resistance network that is detectable in the plasma and spatially validated in tumor tissue. Importantly, a three-marker biomarker panel (LAG3 + HTRA1 + AFP) demonstrated strong discrimination between refractory and treatment-naïve disease (Figure 5D, AUC = 0.821) with good calibration and clinical benefit, suggesting its potential utility for pre-treatment risk stratification.

A key and unexpected finding of the integrated lipidomic analysis was the systematic depletion of sphingolipid and glycerophospholipid species in the refractory plasma. Among the 1,059 lipid species profiled, more than 70% of sphingolipid species were downregulated in refractory patients, with significant depletion of lysophosphatidylserine (LPS), phosphatidylserine (PS), and lysophosphatidylethanolamine (LPE) (Figure 2B). Lysophosphatidylserine (LPS) is a bioactive lysophospholipid known to modulate immune signaling through G protein–coupled receptors, such as GPR34 and related receptors^21^, promoting dendritic cell activation and interferon-γ secretion^22^. Therefore, its depletion represents a potential metabolic mechanism for impaired antitumor immune surveillance. Similarly, phosphatidylserine plays a central role in apoptotic cell recognition and clearance, and reduced PS levels may reflect the suppression of apoptotic signalling pathways in refractory tumors ^23^. These findings were supported by joint pathway enrichment analysis, which identified sphingolipid signalling and metabolism as the most significantly enriched pathways across proteomic and metabolomic datasets (Figure 1D), highlighting the central role of sphingolipid dysregulation in refractory GCT biology.

Consistent with this metabolic reprogramming, the plasma proteome revealed coordinated upregulation of immune checkpoints and stromal remodelling proteins. LAG3 has emerged as one of the most strongly elevated proteins in refractory patients and has been confirmed using ELISA. LAG3 is an inhibitory immune checkpoint receptor expressed on exhausted T cells that suppresses T cell receptor signalling through interactions with MHC class II molecules and the soluble ligand FGL1^24^. Previous studies have demonstrated that LAG3 cooperates with PD-1 to induce T-cell exhaustion in multiple malignancies^25^. In our cohort, LAG3 protein levels correlated strongly with immune pathway activity and the ssGSEA immune score across patients (Supp Fig 2A). Importantly, multiplex immunofluorescence analysis demonstrated that CD8 + LAG3⁺ exhausted T cells were significantly closer to CK-PAN⁺ tumor cells in refractory tumors (Figure 5), indicating a spatial configuration consistent with that of direct immunosuppression. This observation suggests that refractory pediatric GCT do not simply exclude immune cells but instead actively recruit and functionally suppress tumor-reactive T cells.

HTRA1 is the second key effector protein associated with refractory diseases. HTRA1 is a secreted serine protease involved in ECM degradation and regulation of TGF-β signalling^26^. In our dataset, HTRA1 expression correlated strongly with ECM remodelling signatures and immune pathway activity (Supp Fig 2C), positioning it at the intersection of stromal activation and immune suppression. Mechanistically, HTRA1-mediated cleavage of ECM components such as fibronectin and laminin may remodel the tumor microenvironment to facilitate tumor invasion and restrict effective immune infiltration^27,28^. Network analysis further revealed that HTRA1 occupies a central hub position within the integrated multi-omics network, linking lipid metabolic depletion to immune checkpoint activation (Supp Fig 2E). These findings suggest that HTRA1 acts as a molecular bridge connecting metabolic perturbations with microenvironmental remodelling in refractory diseases.

Upstream regulatory analysis identified the suppression of NF-κB and STAT3 inflammatory signalling pathways as a potential mechanistic driver of this resistance program (Figure 3). Although NF-κB is frequently considered a pro-tumorigenic pathway, it also regulates multiple immunostimulatory cytokines and antigen-presentation components^29^. Suppression of NF-κB signalling may therefore contribute to immune evasion by reducing inflammatory signalling and antigen presentation^30^. We propose a model in which sphingolipid depletion disrupts PKC-dependent IKKβ activation, potentially leading to reduced NF-κB signaling activity. This, in turn, represses HTRA1 expression and promotes immune checkpoint activation, culminating in a tumor microenvironment characterized by ECM remodelling and LAG3-mediated T-cell exhaustion.

From a translational perspective, the plasma biomarker panel developed in this study represents a practical clinical application of these findings. AFP alone demonstrated limited discriminatory power, consistent with its known limitations in predicting treatment resistance. In contrast, both HTRA1 and LAG3 individually showed improved predictive performance, and their combination with AFP significantly enhanced the model discrimination (Figure 5D). Decision curve analysis confirmed that the three-marker model provided meaningful clinical benefits across a wide range of risk thresholds (Figure 5H). These results suggest that the integration of immune- and metabolism-related biomarkers may substantially improve early identification of high-risk patients. In addition, the observed enrichment of LAG3-mediated immune exhaustion provides a rationale for exploring LAG3-targeted immunotherapy strategies for refractory pediatric GCT.

Our findings expand the current understanding of GCT biology. Although immune checkpoint pathways have been implicated in adult testicular GCT, plasma-based multi-omics analyses in pediatric patients are largely lacking. Therefore, the present study provides one of the most comprehensive molecular characterizations of refractory pediatric GCT to date and highlights the importance of systemic tumor–immune interactions detectable through liquid biopsy approaches.

This study has several limitations. First, lipidomics analysis was conducted in an exploratory subset due to sample availability, limiting the statistical power for individual lipid species analysis. Second, pathway analyses were derived from plasma proteomics data rather than tumor transcriptomics, and while plasma measurements capture systemic tumor–immune interactions, they may not fully reflect intratumoral signalling activity. Third, the biomarker model was internally validated but required external validation in independent cohorts. Finally, the mechanistic cascade linking sphingolipid depletion, NF-κB suppression, HTRA1 activation, and LAG3-mediated immune exhaustion remains correlated and warrants further functional validation in experimental models.

In conclusion, this integrated multi-omics analysis identifies the sphingolipid–HTRA1–LAG3 axis as a defining feature of platinum-refractory pediatric germ cell tumors. Spatial validation of LAG3-associated immune exhaustion provides mechanistic insights into tumor immune evasion, while the plasma biomarker panel offers a clinically feasible strategy for early risk stratification. Prospective validation across international pediatric oncology cohorts is essential to translate these findings into clinical practice and evaluate therapeutic strategies targeting the LAG3-associated immune exhaustion pathway.

## Conclusion

In summary, this study provides a comprehensive plasma multi-omics characterization of pediatric germ cell tumors and identifies a convergent sphingolipid– NF- κ B– HTRA1/LAG3 regulatory axis associated with treatment resistance. Elevated plasma LAG3 serves as an independent prognostic biomarker and reflects spatially organized immune exhaustion within the tumor microenvironment. Integrated lipidomic and proteomic analyses have further revealed the coordinated disruption of sphingolipid metabolism and inflammatory signaling in refractory diseases. Importantly, a plasma-based biomarker panel integrating LAG3, HTRA1, and AFP levels demonstrated an improved predictive performance for refractory risk. Together, these findings establish a multi-omics framework for risk stratification and suggest potential metabolic-immune therapeutic targets for refractory pediatric germ cell tumors.

## Data Availability

All data produced in the present study are available upon reasonable request to the authors

## Ethics approval and consent to participate

This study was approved by the Ethics Committee of Shandong Cancer Hospital and Institute Affiliated to Shandong First Medical University (approval number: SDTHEC 202508014). All procedures involving human participants were conducted in accordance with the Declaration of Helsinki and relevant institutional guidelines. Written informed consent was obtained from patients or their legal guardians prior to sample collection.

## Consent for publication

Written informed consent for publication was obtained from all participants or their legal guardians.

## Availability of data and materials

The datasets generated and/or analyzed during the current study are available from the corresponding author upon reasonable request.

## Authors’ contributions

Conceptualization: Manfei Liang, Jingfu Wang

Methodology: Yixiang Song, Linke Yang, Guanhua Liu

Data analysis: Qingyou Zhao, Hongtao Li

Investigation: Shuping Yang, Zhongkun Guo, Siying Liu,Zhan Lei

Writing – original draft: Manfei Liang

Writing – review and editing: Jingfu Wang,Qingyou Zhao

Supervision: Manfei Liang

Funding acquisition: Manfei Liang, Yixiang Song

All authors read and approved the final manuscript.

## Acknowledgements

We sincerely thank all patients and their families who participated in this study. We also acknowledge the clinicians and laboratory staff at Shandong First Medical University and collaborating hospitals for their assistance with sample collection and technical support.

## Declaration of generative AI and AI-assisted technologies in the manuscript preparation process

During the preparation of this work, we used Claude in order to the purpose of language refinement and polishing. After using this tool/service, we reviewed and edited the content as needed and take full responsibility for the content of the published article.

## Supplemental Figure legends

Supplementary Figure 1. Model validation and clinical correlation of refractory biomarkers.

(A)Principal component analysis (PCA) of the combined proteomic (green circles) and metabolomic (orange triangles) profiles. The analysis shows concordant separation patterns between the two omics layers for treatment-naïve (light shades) and refractory (dark shades) groups. The gray ellipse represents the 95% confidence interval.

Variable importance analysis. Bar plots show the top 20 most important features for classifying refractory vs. naïve patients. The importance score reflects the contribution of each feature to the model’s predictive accuracy.

(C-D) Plasma protein levels of LAG3 and HTRA1 in patients stratified by serum alpha-fetoprotein (AFP) status. AFP groups were defined using a cutoff of 10 ng/mL (Positive: >10 ng/mL; Negative: <10 ng/mL). The AFP-positive (AFP+) group represents patients with elevated serum AFP levels, a marker associated with certain GCT subtypes like yolk sac tumor (YST). The AFP-negative (AFP-) group includes patients with normal AFP levels, such as those with mature teratoma. Center lines of box plots represent medians, box limits indicate the interquartile range (IQR), whiskers extend to 1.5× IQR, and individual data points are overlaid.

(E-F) Plasma protein levels of LAG3 and HTRA1 in patients with different major histological subtypes: yolk sac tumor (YST) and Others. These subtypes represent distinct biological entities within pediatric GCTs.

Supplementary Figure S2. Pathway activity analysis and multi-omics correlation network associated with the HTRA1–LAG3 regulatory axis.

(A) Gene set enrichment analysis (GSEA) of hallmark and curated biological pathways comparing refractory (RF) and treatment-naïve (NT) tumors. Pathways related to immune signaling, extracellular matrix (ECM) remodeling, lipid metabolism, and inflammatory responses showed significant enrichment in refractory tumors.

(B) Correlation analysis between LAG3 expression and immune pathway activity score estimated by ssGSEA. A strong positive association was observed, suggesting that LAG3 expression reflects increased immune signaling activity in the tumor microenvironment.

(C) Correlation between HTRA1 expression and ECM remodeling pathway activity, indicating that HTRA1 expression is closely linked with extracellular matrix remodeling processes.

(D) Correlation between HTRA1 expression and immune pathway activity score, supporting a functional connection between HTRA1 signaling and immune microenvironment remodeling.

(E) Integrated multi-omics correlation network linking lipid metabolites, protein biomarkers, and pathway activity scores. Lysophospholipids (LPS, LPE, and PS) form a functional module connected to HTRA1, LAG3, ECM remodeling, and immune signaling, suggesting coordinated lipid–immune interactions.

(F) Systemic interaction network highlighting HTRA1 as a central hub protein connecting immune regulators, extracellular matrix components, and lipid-associated pathways.

