## Supplementary material for "Integrated Multi-omics Analysis of 105 Pediatric Germ Cell Tumors Identifies a Sphingolipid – HTRA1 – LAG3 Axis Associated with Immune Evasion in Refractory Disease": Supplementary word.docx

Supplementary Table 1: Detailed Information of Antibodies and ELISA Kits Used in This Study

| **No.** | **Reagent Type** | **Target** | **Manufacturer** | **Catalog No.** | **Host Species** | **Working Concentration** |
| --- | --- | --- | --- | --- | --- | --- |
| 1 | Primary Antibody | HTRA1 | Proteintech | 55011-1-AP | Rabbit | 1:400 |
| 2 | Primary Antibody | LAG3 | Huabio | HA721358 | Rabbit | 1:500 |
| 3 | Primary Antibody | CD8 | AiFang Biologcal | AF20211 | Mouse | 1:500 |
| 4 | Primary Antibody | CD68 | AiFang Biologcal | AF20022 | Mouse | 1:500 |
| 5 | Primary Antibody | CK-PAN | AiFang Biologcal | AF20164 | Mouse | 1:3000 |
| **6** | Secondary Antibody | anti-mouse HRP | AiFang Biologcal | AFIHC001 | Goat | 1:5000 |
| **7** | Secondary Antibody | **anti-rabbit HRP** | AiFang Biologcal | AFIHC002 | Goat | 1:5000 |
| 8 | ELISA Kit | Human HTRA1 ELISA Kit | CUSABIO | CSB-EL010901HU-96 | Human | Operate according to the kit manual; Standard range: 1.56 ng/mL-100 ng/mL |
| 9 | ELISA Kit | Human LAG3 ELISA Kit | CUSABIO | CSB-EL012719HU-96 | Human | Operate according to the kit manual; Standard range: 0.156 ng/mL-10 ng/mL |
