## Supplementary figures and images for "Integrated Multi-omics Analysis of 105 Pediatric Germ Cell Tumors Identifies a Sphingolipid – HTRA1 – LAG3 Axis Associated with Immune Evasion in Refractory Disease"

### Supp Fig S1.tif

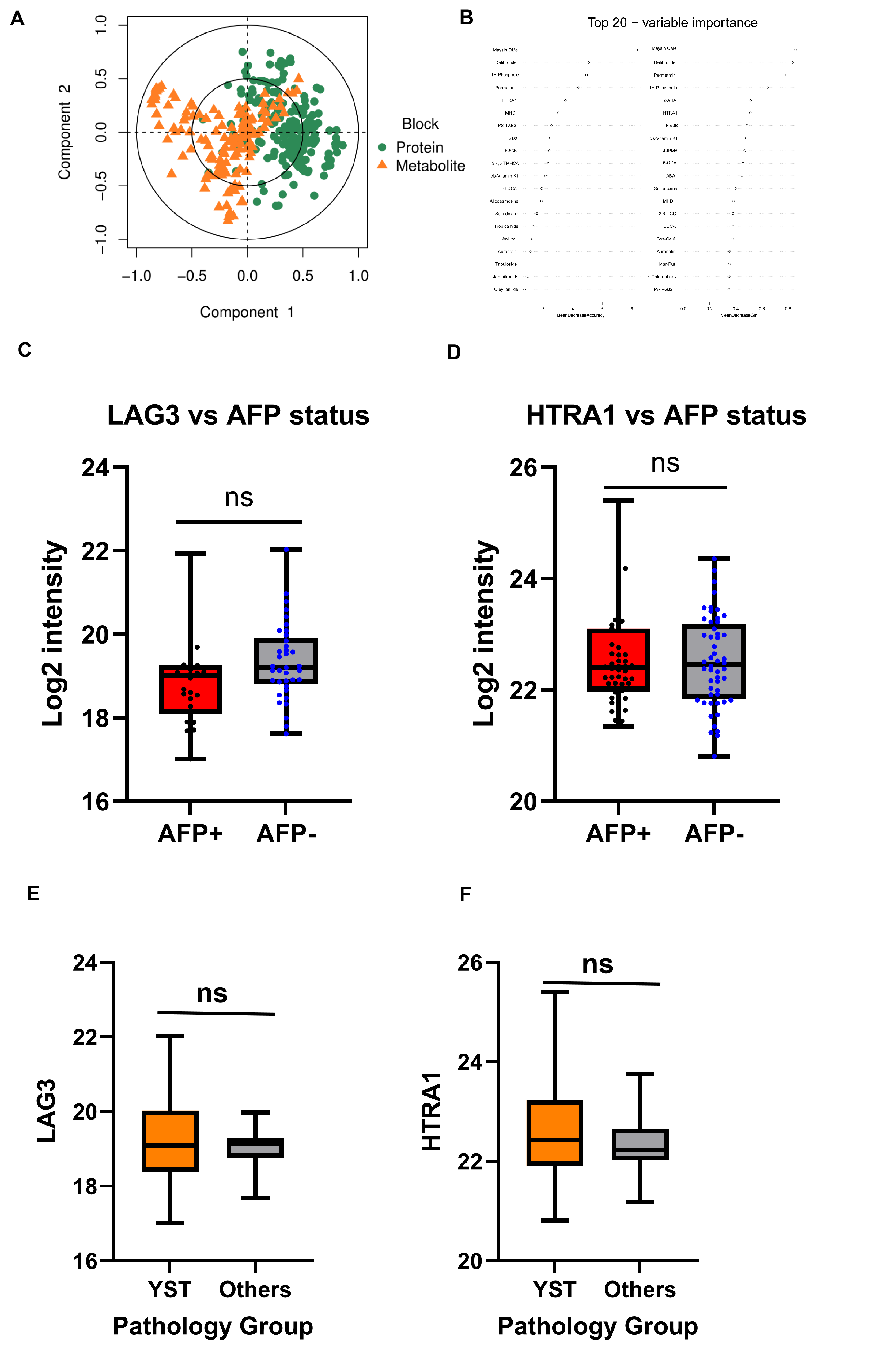

### Supple Fig S2.tif

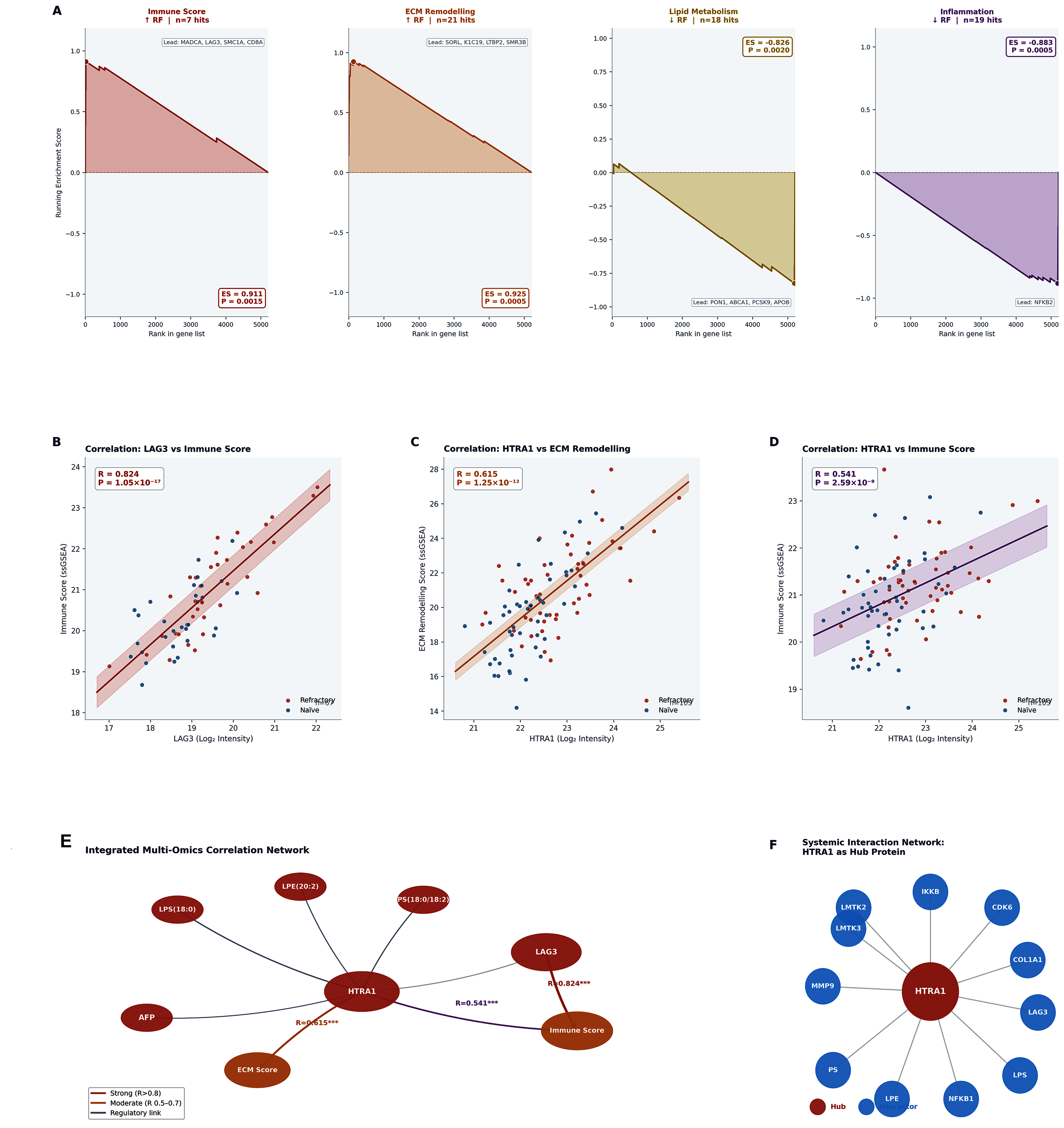
